# Aging as an active player in Alzheimer’s Disease Classification: Insights from feature selection in BrainAge Models

**DOI:** 10.1101/2025.04.16.25325953

**Authors:** Jorge Garcia Condado, Ines Verdugo Recuero, Iñigo Tellaetxe Elorriaga, Colin Birkhenbil, Maria Carrigan, Ibai Diez, Rachel F Buckley, Asier Erramuzpe, Jesus M Cortes, the Alzheimer’s Disease Neuroimaging Initiative

## Abstract

**Background:** BrainAge models estimate the biological age of the brain using neuroimaging or clinical features, making them promising tools for studying neurodegenerative diseases like Alzheimer’s disease. However, the reliance of BrainAge models on neuroimaging features such as grey matter volume and hippocampal atrophy, can introduce biases linked to individuals’ ages as these features are influenced both by normal aging and Alzheimer’s disease progression. The potential presence of such age-biases raises a critical question: can BrainAge models trained to estimate biological brain aging make meaningful contributions to Alzheimer’s diagnosis, or does any introduced age-bias conflate aging effects with disease pathology? Understanding how deliberate feature selection impacts this confounding effect is essential for developing reliable age-related biomarkers.

**Methodology:** We ranked neuroimaging and neuropsychological features based on their mutual information with age and their discriminative power across four clinical groups: cognitively normal, Mild Cognitive Impairment, Alzheimer’s Disease, stable Mild Cognitive Impairment and progressive Mild Cognitive Impairment. Iteratively, we trained BrainAge models using different subsets of these features, some optimized for predicting aging and others for discrimination of clinical Alzheimer’s disease stages. We assess the error in BrainAge delta, the difference between predicted biological age and chronological age, and evaluate its classification performance across clinical groups. Finally, we compare using deltas for classification with a logistic regression model directly trained on the neuroimaging features used in the BrainAge models.

**Results:** Neuroimaging features are more strongly correlated with aging, while neuropsychological features exhibit greater discriminative power for Alzheimer’s disease classification. BrainAge models optimized for age prediction yield deltas that are suboptimal when used for classifying Alzheimeŕs disease, whereas models trained to generate deltas optimized to be used for classifying Alzheimer’s disease have reduced age prediction accuracy. This trade-off suggests that BrainAge models may not optimally separate aging-related changes from disease-specific alterations. BrainAge models have varying classification accuracy as compared to direct utilization of features in logistic regression. However, BrainAge provides a continuous measure, offering a single output that can be used across clinical stages, in contrast to classification approaches that require explicit labels for each disease stage.

**Conclusion:** Aging significantly affects BrainAge-based classification of Alzheimer’s disease. Feature selection plays a critical role in mitigating this effect, as the outputs of models trained to predict age, the deltas, may fail in Alzheimeŕs disease classification. These findings underscore the need for task-specific feature selection and model design to ensure that BrainAge models are appropriately applied in neurodegenerative disease research.

## 1. Introduction

BrainAge models are predictive tools developed using machine learning or deep learning techniques to estimate an individual’s chronological age based on neuroimaging data (Franke et al., 2010). BrainAge models are often employed to assess deviations from typical aging trajectories, with the difference between a participant’s predicted and actual age, referred to as the BrainAge delta, serving as a potential biomarker for various diseases. A positive delta indicates that the brain appears older than expected in reference to a healthy brain of that same age, whereas a negative delta suggests a younger-appearing brain. This metric is increasingly used as a biomarker of brain health, with higher BrainAge deltas linked to neurodegeneration and cognitive decline while lower deltas may reflect resilience to aging processes. These models have gained significant attention in the study of neurodegenerative diseases, such as Alzheimer’s disease (AD) (Gaser et al., 2013) as well as psychiatric disorders and other neurological conditions (Baecker et al., 2021).

Despite the potential of BrainAge models, several challenges hinder their generalizability and clinical utility. One critical issue is the inherent bias introduced by training models on sample populations that are not representative of the broader population. For instance, most models are developed using data from predominantly white individuals, which can result in biased predictions when applied to diverse populations (Moguilner et al., 2024; Puc et al., 2021). Additionally, the definition of “healthy” in age modeling is varies depending on the study, and this variability in training data can further skew results (de Lange et al., 2022; Franke and Gaser, 2019).

Another major challenge is the reproducibility of neuroimaging-based models, especially across different imaging sites and protocols (Korbmacher et al., 2023; Yu et al., 2024). Variability in image acquisition, preprocessing, and analysis methods can introduce noise, complicating efforts to develop models that are robust and generalizable (Gebre et al., 2023). Although initiatives like the Brain Age Standardized Evaluation (BASE) (Dular and Špiclin, 2024) aim to standardize BrainAge prediction workflows, there remains a need for consensus on evaluation criteria and the consistent application of these standards.

Our previous work has demonstrated that BrainAge models with poorer age prediction accuracy can, at times, yield deltas that perform better when used as input in downstream tasks, such as classifying between stable Mild Cognitive Impairment (sMCI) and progressive MCI (pMCI) to AD (Garcia Condado et al., 2023). This suggests that optimizing models purely for age prediction may not necessarily lead to better outcomes when using the deltas to distinguish between individuals with AD dementia and cognitively normal individuals. Despite aging being a strong predictor of dementia, it is non-specific to any one disease, making it challenging to distinguish between normal and pathological ageing.

In this work, we aim to explore how the choice of features used in BrainAge models affects both the error in age prediction and the performance of deltas when used as input in auxiliary downstream tasks, such as classifying between clinical groups. Specifically, we investigate these relationships in the context of AD to elucidate the trade-offs between prediction accuracy and classification performance. We rank neuroimaging and neuropsychological features based on their relationship with age and their discriminative power across clinical groups. By constructing BrainAge models with different feature sets, we assess the error in BrainAge delta and its classification performance when used as input to a logistic regressor. Finally, we compare the predictive value of BrainAge delta to a direct classification approach using neuroimaging features.

## 2. Materials and Methods

In this study, we explore two different approaches to BrainAge modeling, see Fig. 1. Our first approach explores how BrainAge models trained on different feature sets compare in their age prediction error compared to the use of the generated deltas in the downstream task of classifying two different clinical groups. Our second approach explores whether using the BrainAge deltas for classification is better than using the features directly with a logistic regression for classification.

**Figure 1.**
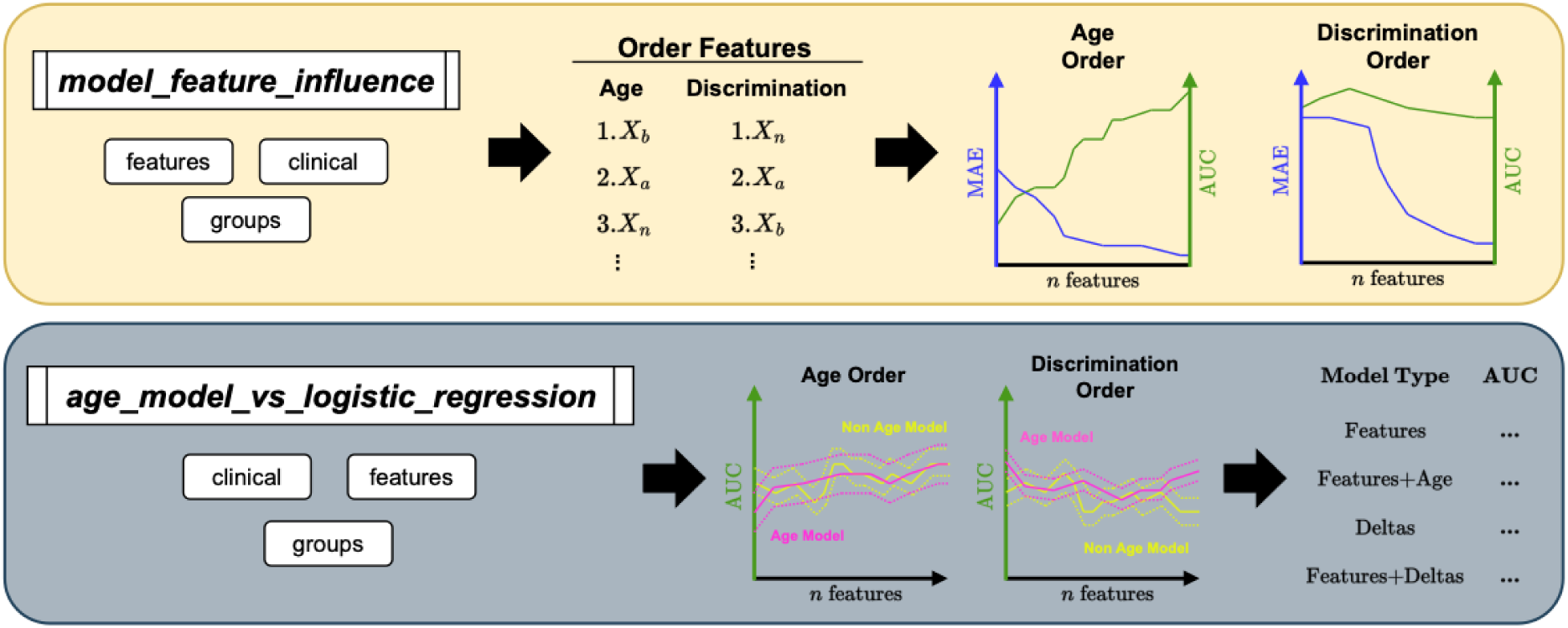
Overview of the two new commands for AgeML to explore the relationship between age prediction and classification accuracy. *Top panel:* Model Feature Influence pipeline schematic. First, the provided features are ranked according to their mutual information with age or according to their discriminative power to classify the specified clinical groups. Then, age regression and clinical classification models are trained with the computed orderings to evaluate how the progressive addition of features affects the performance of the models. The progression curves are automatically plotted. *Bottom panel:* Age Models versus direct logistic regression pipeline. The given features are first ranked according to the same criterion from above. After, two logistic regressor types are trained to classify the specified clinical groups; one based on an age delta computed from Age models; and the other directly using the features. The performance of the classifiers is plotted trained with increasingly more features added in the computed orderings, similar to the panel above. A summary table and the progression curves are automatically output.

Both methodologies have been integrated for easy use within AgeML (Garcia Condado et al., 2025). AgeML is an OpenSource Python package for Age Modeling with Machine Learning. The first approach is integrated under the command ***model_feature_influence***, see Fig. 1 Top panel. The second approach is integrated under the command ***age_model_vs_logistic_regression***, see Fig. 1 Bottom panel. The OpenSource code can be found in the GitHub repository: github.com/compneurobilbao/ageml.

### 2.1 Data

Data used in the preparation of this article was obtained from the Alzheimer’s Disease Neuroimaging Initiative (ADNI) database (adni.loni.usc.edu) (Petersen et al., 2010). All participants in the ADNI2 and ADNI3 phases who had an initial visit with T1-weighted imaging and neuropsychological evaluation were extracted from the ADNI database. This included clinically normal older adults (CN), and MCI and AD participants. The demographics are provided in Table 1.

**Table 1.** Cohort demographics.

| Characteristic | CN<br>N = 629<sup>1</sup> | MCI<br>N = 635<sup>1</sup> | AD<br>N = 208<sup>1</sup> | sMCI<br>N = 238<sup>1</sup> | pMCI<br>N = 98<sup>1</sup> | p-value<sup>2</sup> |
| --- | --- | --- | --- | --- | --- | --- |
| Gender |  |  |  |  |  | <0.001 |
| Female | 364 (58%) | 279 (44%) | 87 (42%) | 110 (46%) | 43 (44%) |  |
| Male | 265 (42%) | 356 (56%) | 121 (58%) | 128 (54%) | 55 (56%) |  |
| Age (Years) | 72 (7) | 72 (8) | 75 (8) | 73 (8) | 73 (7) | <0.001 |
| Education (Years) | 17 (2) | 16 (3) | 16 (3) | 16 (3) | 16 (3) | <0.001 |

<sup>1</sup>n (%); Mean (SD)
<sup>2</sup>Pearson's Chi-squared test; Kruskal-Wallis rank sum test

Using longitudinal data, we identified those who progressed from MCI to AD. sMCI participants were considered stable if they continued to be diagnosed with MCI after 3 years from the initial visit. Similarly, only participants who progressed to AD within 3 years from the initial visit were considered pMCI.

Models were built using structural brain features extracted from T1-weighted images and neuropsychological features at baseline. Ten neuroimaging volumetric features were obtained from T1-weighted images from each participant’s first visit. Three of the ten features were extracted with the Structural Image Evaluation with Normalisation of Atrophy Cross-sectional (SIENAX) (Smith et al., 2002), part of FMRIB Software Library (FSL) (Smith et al., 2004) to obtain grey matter volume, white matter volume, cerebrospinal fluid volume, as well as a volume scale value. Then the other seven features were extracted using the FMRIB’s Integrated Registration and Segmentation Tool (FIRST) (Patenaude et al., 2011) to segment and calculate the volumes of the thalamus, caudate, putamen, pallidum, hippocampus, amygdala, and accumbens over both hemispheres. The volume scale value was used to control for differences in brain size. We also extracted these same features using FreeSurfer 7.2 (Fischl, 2012) and cortical thickness measurements for the inferior parietal, inferior temporal, middle temporal, entorhinal, parahippocampal and fusiform regions. Accuracy of the segmentation tools, both sensitivity and specificity, are published elsewhere (Fischl, 2012; Patenaude et al., 2011; Smith et al., 2002). We selected features from T1-weighted imaging due to their lower variability in signal-to-noise ratio and reduced site-related differences (Warrington et al., 2025), as well as prior evidence that structural metrics outperform functional MRI measures in age modeling (Guan et al., 2023). In particular, we focused on subcortical volumes and cortical thickness because they are strongly implicated in both aging and Alzheimer’s disease such as the hippocampus (Sabuncu, 2011), amygdala (Poulin et al., 2011) and cortical thinning of the cortex (Bakkour et al., 2009).

Six neuropsychological features were obtained from neuropsychological assessments from each participant’s first visit. These consisted of scores from standard neuropsychological tests: Mini-Mental State Examination (MMSE)(Folstein et al., 1975), Alzheimer’s Disease Assessment Scale (ADAS) (“A new rating scale for Alzheimer’s disease,” 1984), Functional Assessment Questionaire (FAQ) (Pfeffer et al., 1982), and Montreal Cognitive Assessment (MoCA) (Nasreddine et al., 2005), as well as two metrics generated in the ADNI study: ADNI Memory score (for the Alzheimer’s Disease Neuroimaging Initiative et al., 2012a) and ADNI Executive Function (for the Alzheimer’s Disease Neuroimaging Initiative et al., 2012b). We compare 4 different clinical groups in the study: CN vs MCI, CN vs AD, MCI vs AD and sMCI vs pMCI.

### 2.2 Age prediction and classification accuracy for different feature sets

To create the different feature sets, we first order the features based on mutual information (MI), which is well known for capturing relationships beyond the linear dependencies detected by pairwise correlations. In particular, defined from information theory, MI quantifies the statistical dependency between variables, providing a measure of how much knowing one variable reduces uncertainty about another. We use MI to rank features according to two criteria. First, features are ordered by their MI with age using the function *mutual_information_regression* from sklearn (“Scikit-learn: Machine Learning in Python,” n.d.). This ranking reflects the extent to which each feature individually correlates with age. Second, features are ranked based on their discriminative power between two clinical groups by calculating their MI with clinical labels using the function *mutual_information_classif* from sklearn (“Scikit-learn: Machine Learning in Python,” n.d.). This approach identifies features most relevant for distinguishing between clinical labels. For each different case, feature sets are created by starting with the feature with the highest mutual information and iteratively adding the next feature with the next highest mutual information. In this study, we use 10 neuroimaging features (grey matter, white matter, cerebrospinal fluid, thalamus, caudate, putamen, pallidum, hippocampus, amygdala, and accumbens normalized volumes) and 6 neuropsychological features (MMSE, ADAS, FAQ, MoCA, ADNI Memory and ADNI executive), so a total of 16 feature sets are created.

For each feature set, we train a BrainAge model based on healthy controls. Specific details on the model training can be found in the Methods section of AgeML (Garcia Condado et al., 2025). In summary, with *y* as age, ***X*** as our feature set and *f*() as our pipeline, we optimize the parameters of our pipeline *f*() by means of minimizing the mean squared error of age on cognitively normal controls. Our pipeline consists of a feature scaler and a linear regressor and we use a 5-fold cross-validation scheme. The reported age prediction error, the Mean Absolute Error (MAE), is reported before the age bias correction step (de Lange and Cole, 2020).

Afterwards we apply the BrainAge model to the two clinical groups of interest to obtain predicted ages. We then calculate age deltas, the predicted age after age bias correction minus the chronological age, for each participant. If one of the clinical groups is the control group, the predicted ages from the cross-validation out-of-fold predictions are used. Then the deltas are used as input into a logistic regressor to classify between two groups and obtain Areas Under the Curve (AUC). To handle class imbalance within cross-validation, we first stratify the n-fold CV split to preserve the original class ratios. Within each training fold, we then apply undersampling of the majority class to match the number of participants in the smaller group. This procedure prevents information leakage whilst the test fold remains representative of the original population distribution. This avoids optimistic bias because class rebalancing was carried out separately within each cross-validation fold before computing the AUC. To address potential circularity in using cognitive test to build BrainAge models whose deltas are later used for classification, we trained the BrainAge model exclusively on healthy controls, preventing data leakage from MCI or AD subjects as argued in previous studies (Garcia Condado et al., 2023).

For each BrainAge model trained on each feature set, we obtain a MAE for the age prediction task and an AUC from using the deltas in the downstream classification task. We then plot the MAE and AUC over the number of features used in training for each model and repeat this process for the two different types of ordering.

### 2.3 Classification accuracy for different models

We also examine how classification accuracy varies across different models. Specifically, we train four logistic regression models using different inputs. Three models use BrainAge deltas as input to a logistic regressor, where the deltas are derived from three different BrainAge models: a linear regressor, a Ridge regressor, and an SVM. In addition, we train a separate logistic regression model directly on the feature sets. For this analysis, only neuroimaging features are used, as some neuropsychological test features may have been considered by clinicians when assigning clinical labels. Excluding these features helps prevent potential biases in classification when using directly the features in classification.

We are also interested in understanding the value of the BrainAge deltas in terms of whether it adds extra information to the classification task. Therefore, we also train 4 distinct logistic regressors and compare their AUC. The 4 logistic regressors use the following as input: all neuroimaging features, all neuroimaging features and the age of participants, only the BrainAge delta, all neuroimaging features and the BrainAge delta.

## 3. Results

### 3.1 Age prediction error and classification accuracy for different feature sets

The results of the feature ranking are shown in Table 2, listed in descending order of importance. The “Age” column refers to the relationship with age, while the remaining columns reflect the discriminatory power of each feature between clinical groups. We look at discrimination between 4 scenarios: CN vs AD, CN vs MCI, MCI vs AD and sMCI vs pMCI. Neuroimaging features were found to be more informative for inferring age, whereas neuropsychological features proved more effective in discriminating between clinical groups. When mapping MI with age and the discriminative power of each feature in Fig. 2, neuroimaging features show higher MI with age than with classification, whereas neuropsychological features show lower MI with age than with classification. However, neuropsychological features also show low MI when attempting to distinguish between sMCI and pMCI because at baseline these two subgroups are both characterized as MCI and hence, share similar cognitive profiles. Supplementary Fig. 1 shows the correlation between FIRST/FAST segmentation outputs and FreeSurfer outputs. In Supplementary Fig. 2 and Supplementary Table 1 it can be seen that cortical thickness measurements are worse at predicting age than subcortical volumes and are worse than neuropsychological features to distinguish between different groups. However, the entorhinal cortical thickness measurements show greater power at discrimination between CN and AD than neuroimaging features of subcortical volumes. Many of the cortical thickness measurements are also more powerful at discriminating sMCI vs pMCI than subcortical volumes.

**Figure 2.**
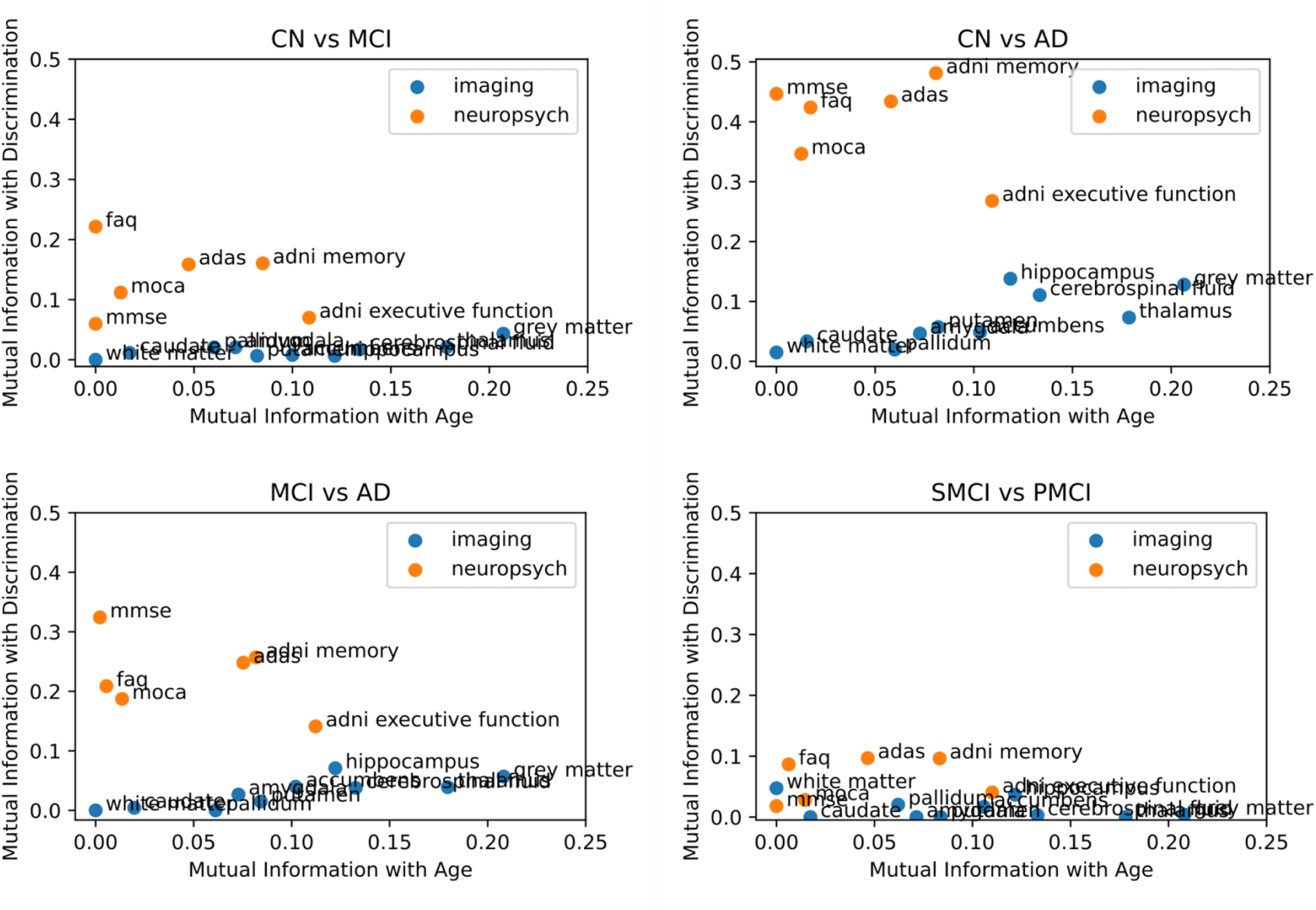
Mutual information of each feature with Age and their discriminative power. In blue are features derived from neuroimaging metrics and in orange features derived from neuropsychological testis. Control (CN), Alzheimer’s Disease (AD), Mild Cognitive Impairment (MCI), stable MCI (sMCI), and progressive MCI (pMCI)

**Table 2.** Results of ranking features across different orderings. The features are ordered in descending importance according to the variable indicated in the column header. The first column indicates ordering according to mutual information with age, while the subsequent columns make a comparison between the mutual information of features for the following groups: Control Group (CN), Alzheimer’s Diseases (AD), Mild Cognitive Impairment (MCI), Stable Mild Cognitive Impairment (sMCI) and Progressive Mild Cognitive Impairment (pMCI).

| Set | Age | CN vs AD | CN vs MCI | MCI vs AD | sMCI vs pMCI |
| --- | --- | --- | --- | --- | --- |
| 1 | Grey Matter | ADNI Memory | ADNI Memory | MMSE | ADAS |
| 2 | Thalamus | MMSE | FAQ | ADAS | ADNI Memory |
| 3 | Cerebrospinal Fluid | FAQ | ADAS | ADNI Memory | MMSE |
| 4 | Hippocampus | ADAS | MMSE | FAQ | FAQ |
| 5 | ADNI Executive Function | MoCA | MoCA | MoCA | White Matter |
| 6 | Accumbens | ADNI Executive Function | ADNI Executive Function | ADNI Executive Function | MoCA |
| 7 | ADNI Memory | Hippocampus | Grey Matter | Hippocampus | ADNI Executive Function |
| 8 | Putamen | Grey Matter | Thalamus | Grey Matter | Hippocampus |
| 9 | Amygdala | Cerebrospinal Fluid | Amygdala | Accumbens | Pallidum |
| 10 | Pallidum | Thalamus | Pallidum | Thalamus | Accumbens |
| 11 | ADAS | Putamen | Cerebrospinal Fluid | Cerebrospinal Fluid | Grey Matter |
| 12 | Caudate | Accumbens | Caudate | Amygdala | Cerebrospinal Fluid |
| 13 | MoCA | Amygdala | Accumbens | Putamen | Thalamus |
| 14 | FAQ | Caudate | Putamen | Caudate | Caudate |
| 15 | White Matter | Pallidum | Hippocampus | White Matter | Putamen |
| 16 | MMSE | White Matter | White Matter | Pallidum | Amygdala |

After obtaining the different feature rankings, the pipeline is executed four times, once for each of the four classification tasks of interest: CN vs AD, CN vs MCI, MCI vs AD and sMCI vs pMCI. Results are shown in Fig. 3. This figure illustrates how MAE and AUC evolve as more features are added to each set, depending on whether the features are more relevant for age modeling or for classifying clinical groups. When features are ranked by their relevance to age, the initial models trained on the top-ranked subset tend to predict age more accurately, but the deltas perform worse in classification tasks. We see the opposite effect when the features are ranked in importance to discriminate. However, when classifying sMCI and pMCI, we observe that the age-based and discrimination-based models begin to overlap earlier as more features are added because neuropsychological features show MI values comparable to those of neuroimaging features in distinguishing between these two groups. In Supplementary Fig. 3 we see that adding cortical thickness measurements yields similar results.

**Figure. 3.**
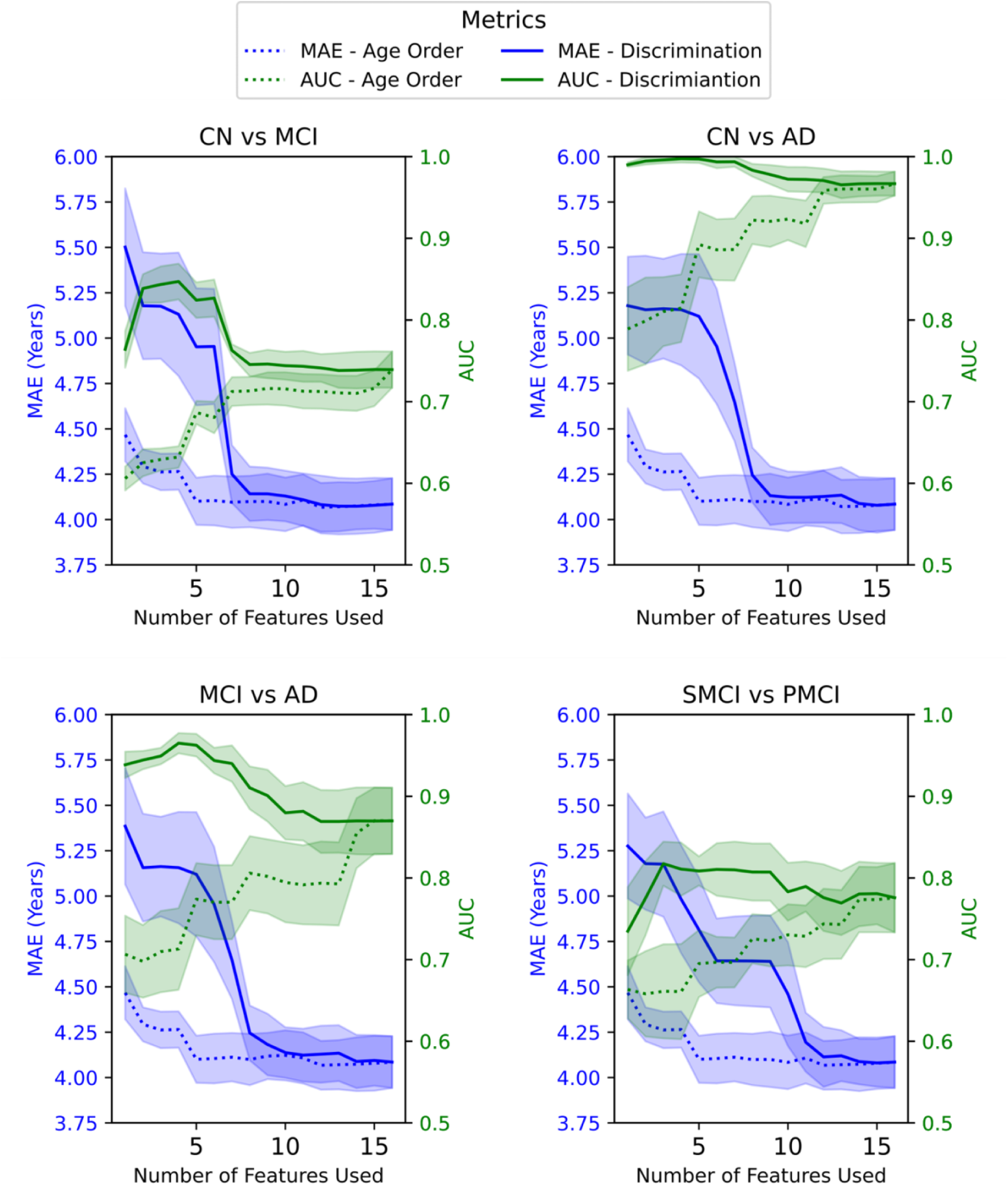
Comparison of the performance metrics, Mean Absolute Error (MAE), and Area Under the Curve (AUC) for different health condition groups using BrainAge Modelling. Features are added to the BrainAge model in descending order based on their age relationship (dotted line). Additionally, features are added to the BrainAge model in descending order according to their importance in discriminating between the following groups: Control (CN), Alzheimer’s Disease (AD), Mild Cognitive Impairment (MCI), stable MCI (sMCI), and progressive MCI (pMCI) (solid line). The shaded areas show the 95% confidence intervals of the MAE and AUC measurements.

### 3.2 Logistic Regression using features vs BrainAge deltas

We trained three different BrainAge models: a linear regressor, a ridge regressor and a SVM with different neuroimaging features. We compared the classification performance of BrainAge deltas with that of using the original features directly in a logistic regressor. The results for all four scenarios --CN vs AD, CN vs MCI, MCI vs AD and sMCI vs pMCI--are shown in Fig. 4. Using the original features as input to the logistic regressor yielded the best performance in CN vs AD and CN vs MCI, similar performance to the other models in MCI vs AD but worse in sMCI vs pMCI. In Supplementary Fig. 4 we see that adding cortical thickness measurements which have higher power of discrimination cause the AUC performance of the logistic regressor trained on the original features to outperform all BrainAge models, that do not see an increase in AUC performance.

**Figure. 4.**
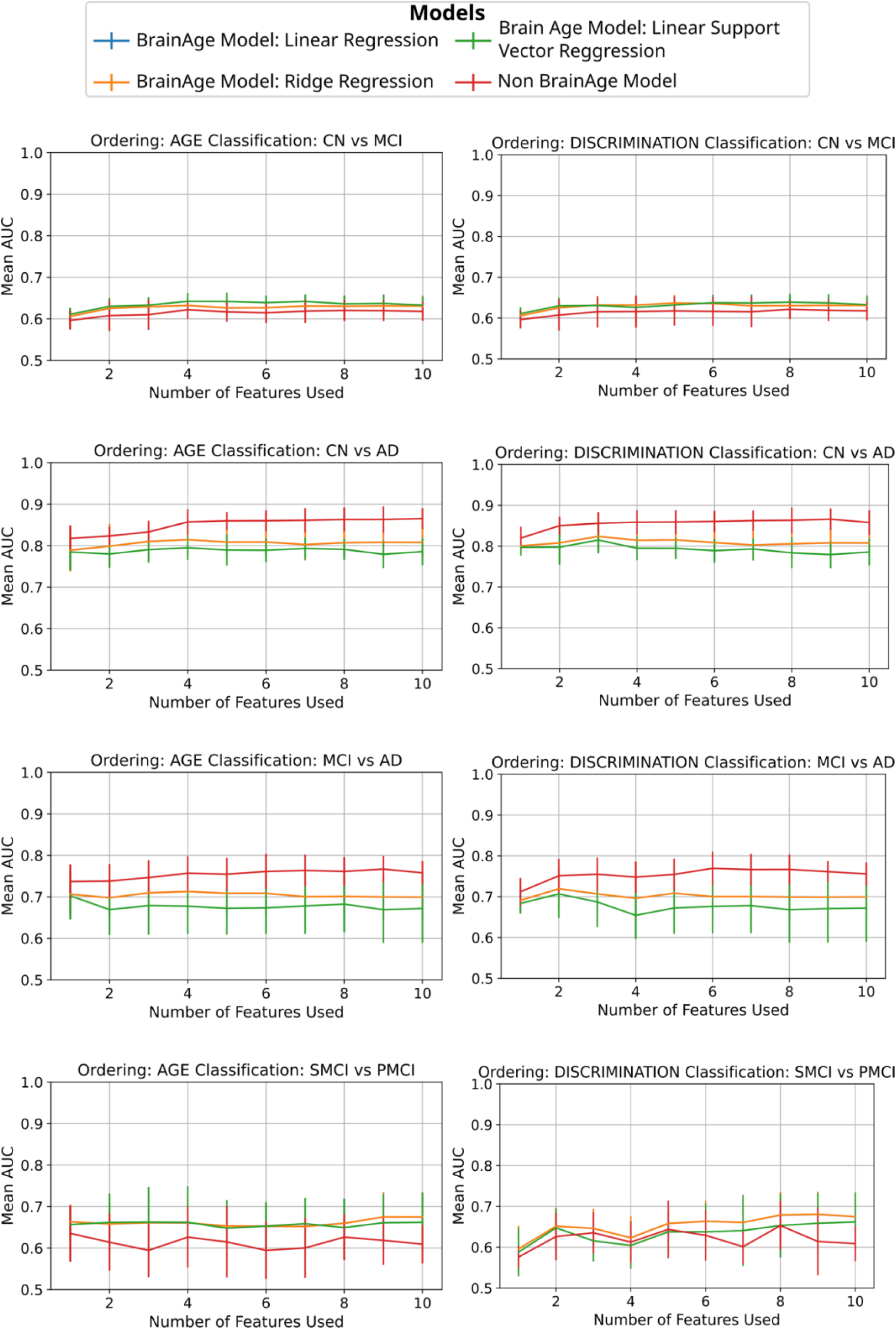
Comparison of the Area Under the Curve (AUC) using different machine learning models for classification using neuroimaging features. *Blue line:* Input to logistic regressor: Delta. BrainAge model: linear regression. *Orange line:* Input to logistic regressor: Delta. BrainAge model: Ridge. *Green line:* Input to logistic regressor: Delta. BrainAge model: Support Vector Regressor. *Red line:* Input to logistic regressor: Neuroimaging Features, No BrainAge modeling. Models are tested and trained across clinical classification groups and features ordering by Age Relationship and by discrimination between groups order (Control Group (CN), Alzheimer Disease (AD), Mild Cognitive Impairment (MCI), stable Mild Cognitive Impairment (sMCI) and progressive Mild Cognitive Impairment (pMCI)). The blue and orange line overlap. Error bars show the standard deviation of the AUC across CV folds.

Finally, using all neuroimaging features, we evaluated whether adding age or the delta as an additional feature could improve classification performance. The results for each scenario are summarized in Table 3. Adding neither age nor the delta appeared to improve the AUC, as most values remained within one standard deviation of each other. Repeating the experiment with 5 different random seeds did not yield fluctuating results as seen in the standard deviations of Supplementary Table 2.

**Table 3.** Comparison of the Area Under the Curve (AUC) across various clinical classifications using different input feature sets. The classification was performed using a Logistic Regressor and included all brain structural features for the following groups: Control Group (CN), Alzheimer’s Disease (AD), Mild Cognitive Impairment (MCI), stable Mild Cognitive Impairment (sMCI), and progressive Mild Cognitive Impairment (pMCI). ± indicates standard deviations across CV folds.

| **Groups** | **Features** | **Features + Age** | **Delta** | **Features + Delta** |
| --- | --- | --- | --- | --- |
| CN vs MCI | $0.62 \pm 0.02$ | $0.63 \pm 0.02$ | $0.63 \pm 0.03$ | $0.63 \pm 0.02$ |
| CN vs AD | $0.86 \pm 0.03$ | $0.87 \pm 0.03$ | $0.81 \pm 0.04$ | $0.87 \pm 0.03$ |
| MCI vs AD | $0.76 \pm 0.03$ | $0.76 \pm 0.03$ | $0.69 \pm 0.05$ | $0.76 \pm 0.04$ |
| pMCI vs sMCI | $0.58 \pm 0.09$ | $0.62 \pm 0.07$ | $0.67 \pm 0.04$ | $0.63 \pm 0.07$ |

## 4. Discussion

This study aimed to evaluate how different neuroimaging and neuropsychological features influence BrainAge predictions and the utility of BrainAge deltas in classifying AD participants. By analyzing the relationship between these features and both age estimation and clinical group distinctions, we assessed the reliability of BrainAge deltas as biomarkers. Our findings provide insight into the strengths and limitations of using BrainAge models for AD classification, highlighting key factors that impact their performance and the need for careful feature selection based on specific research and clinical objectives.

As expected, ranking features by their age prediction accuracy and discriminative power highlights the distinct contributions of neuroimaging and neuropsychological features. Neuroimaging features, specifically grey matter and particularly those related to brain structures like the hippocampus, thalamus, and amygdala, are highly effective in predicting age. In contrast, neuropsychological scores excel at discriminating between clinical groups. This supports the idea that age prediction relies more on the structural integrity of brain regions, while cognitive performance metrics are more sensitive to the pathological distinctions between clinical groups.

It is important to consider possible circularity in using BrainAge deltas built with neuropsychological testing features for classification due to their role in clinical diagnosis, i.e., assigning healthy control, MCI, and AD labels in the first place. In ADNI for example, diagnostic assessment of AD staging includes a selection of these tests in combination with cutoff points and other clinical assessments, such as the Clincal Dementia Rating score not used in this study (Petersen et al., 2010). In the present study, this concern was accounted for by training our BrainAge model on healthy controls and, and thus preventing data leakage in terms of biases in the BrainAge deltas, as the model is not trained on MCI or AD. In the specific case of classifying sMCI and pMCI, despite neuropsychological tests contributing to initial diagnostic classification, our approach avoids circularity by use of baseline scores to predict future conversion rather than reclassifying subjects based on later assessments (Garcia Condado et al., 2023). Excluding neuropsychological features reduced sMCI vs. pMCI classification performance from 0.91 to 0.68 AUC (Garcia Condado et al., 2023). In Fig. 3, this is further illustrated at x = 1, where the full green line is always a neuropsychological feature and achieves a substantially higher AUC than the dotted green line which is a single neuroimaging feature. These findings indicate that neuropsychological tests carry predictive information beyond their traditional role in diagnostic labeling based on cutoffs, supporting their validity for assessing and monitoring disease progression. We acknowledge a potential limitation and bias in using neuropsychological tests that were part of the clinical decision-making process. Further research is needed using more deeply phenotyped cohorts to explore the effect of training on different neuropsychological tests that were not used in the diagnosis labelling process.

Notably, the hippocampus is consistently identified for its high discriminative power across all clinical groups. The hippocampus is among the earliest regions affected by AD pathology, and its atrophy is strongly associated to memory deficits, a hallmark of the disease. In the discrimination tasks, hippocampal volumes are effective at distinguishing between CN, MCI, and AD groups, which aligns with the well-established literature on the role of the hippocampus in cognitive decline (Mu and Gage, 2011). The early inclusion of hippocampal features in both age and discrimination-ordered sets further underscores its dual importance in aging and AD progression. This is consistent with previous findings showing that hippocampal connectivity is indeed significantly affected by aging, but, other circuits—such as the fronto-striato-thalamic network—are even more severely disrupted (see Fig. S4 in Bonifazi et al) (Bonifazi et al., 2018). Thus, although the hippocampus is involved in both aging and AD, its prominent role in AD disease may reflect a stronger association with pathological processes rather than normal aging, highlighting the interaction between these two mechanisms.

Our results show clear distinctions between BrainAge models trained on feature sets ranked by age than those ranked by their discriminative power across clinical groups. When features are ranked based on their relationship with age, the MAE of BrainAge models begin at a lower value, indicating that the model achieves higher age prediction accuracy with the top-ranked features. In this case, when using the produced delta for classification the AUC increases steadily as more features are included in the BrainAge model. Thus, reflecting progressively improved ability of the generated deltas to discriminate between clinical groups. This suggests that age-related features provide a solid foundation for accurate age prediction, while classification performance improves progressively as additional, more diverse features are incorporated.

Conversely, when features are ranked by their discriminative power, the BrainAge model’s delta AUC shows a higher intercept, reflecting better initial classification performance. However, the MAE of the BrainAge model has a higher intercept compared to the age-ranked feature sets, indicating less accurate age predictions at the onset. Hence, better age prediction does not always mean better classification power. A key transition point occurs when neuroimaging features start to appear in the discrimination-ordered set, after which the AUC stabilizes and MAE decreases. However, we do not see an increase in AUC. This indicates that neuroimaging features enhance age prediction but worsen classification accuracy.

Our previous study showed that models just trained on neuropsychological data outperformed those trained on neuroimaging data on the classification task but not in age prediction error (Garcia Condado et al., 2023). We acknowledge that this is to be expected since neuropsychological testing is used to diagnose and is an inherent part of the process. However, as argued above this is not circular reasoning since the BrainAge models are trained in the healthy cohort and never use neuropsychological data from MCI or AD during training. Hence, it is important to determine what the use of the calculated deltas will be when creating BrainAge models. Subsequently, the model should be trained with the features required for its respective task.

Recent studies have emphasized the importance of distinguishing between brain changes due to typical aging and those caused by AD. Hwang et al. (2022) introduced machine learning models such as SPARE-AD and SPARE-BA to decouple the effects of aging and neurodegeneration in brain imaging data (Hwang et al., 2022). Their results showed that by employing conservative molecular diagnoses and introducing Alzheimer’s continuum cases, it was possible to derive more specific neuroanatomical biomarkers for aging and AD, reducing the overlap in brain regions affected by both processes. Similarly, Binette et al. (2020) investigated grey matter differences across a wide age range and in AD patients, finding that while both aging and AD contribute to widespread brain atrophy, AD uniquely disrupts whole-brain morphometric organization (Pichet Binette et al., 2020). This disruption in grey matter pattern, more than volume loss, was strongly associated with cognitive impairment, highlighting the need to account for these distinct mechanisms when assessing BrainAge. Alternatively, feature importance for biological aging FIBA has been proposed as a method to improve the relevance of age models by identifying features that contribute specifically to biological rather than chronological age, refining their association with cognitive traits (Oh et al., 2023).

Our results indicate that training a logistic regression directly on the features either outperforms or equals a logistic regression trained on BrainAge deltas. The results tend to be similar or worse when the classification task is harder, like when distinguishing sMCI vs pMCI. However, when cortical thickness measurements are included, BrainAge models perform worse at distinguishing between sMCI vs pMCI. While a direct classifier can exploit the discriminative value of cortical thickness, this information appears to be lost in the BrainAge modelling process. This is in line with a recent study that found that neural network-based BrainAge models, when retrained for classification, perform worse than models trained directly for classification (Tan et al., 2025). Training on different regression models, such as linear regression ridge regression, and SVM does not improve performance. This outcome is likely influenced by the high linear correlation among neuroimaging features, as a model using a single key feature often performs similarly to those incorporating multiple features. Grey matter volume in particular plays a dominant role in both age prediction and discrimination across clinical groups, highlighting its central importance in distinguishing disease states.

However, BrainAge offers an advantage over training a logistic regressor directly with features. BrainAge deltas encapsulate disease progression within a single metric rather than requiring training separate classification models for each clinical group. Prior studies have shown that BrainAge deltas correlate with the time to conversion from MCI to AD (Garcia Condado et al., 2023), making them a potentially useful quantitative biomarker for assessing risk. This further underscores the need for careful consideration when applying BrainAge models to clinical decision-making.

Adding age or BrainAge delta as additional features does not consistently improve classification accuracy, as indicated by minimal AUC increases that remain within standard deviations. This variability suggests that neither age nor delta provides substantial independent discriminative value beyond the primary neuroimaging features. Notably, repeating the classification process with different data partitions and cross-validation folds leads to wide fluctuations in results, emphasizing the sensitivity of the models to sample variation. This finding points to the need for further research to better understand the potential utility of age and delta features in classification, as well as the robustness of these models across varying sample conditions. Recent studies suggest that using chronological age as a pretraining target might be suboptimal for predicting specific health outcomes (Tan et al., 2025).

A limitation of this work is that only neuroimaging features from a single modality, T1-weighted images, are included in the analysis. Features from these modalities were chosen because there is lower variability compared to diffusion or functional brain imaging (Warrington et al., 2025). While the ADNI dataset offers rich multimodal imaging, it was collected across a large number of scanners with relatively few subjects per site (e.g., fewer than 10 participants in many centers), which introduces substantial site-related variability. This effect is particularly problematic for modalities such as diffusion MRI and functional MRI, where sequence parameters, scanner hardware, and acquisition protocols strongly influence signal-to-noise ratios and overall reproducibility. Previous studies have also shown that structural based metrics outperform functional MRI metrics at the age modelling prediction task (Guan et al., 2023). Including more quantitative measurements such as cortical thickness measurements did not improve age prediction accuracies in line with previous studies (Guan et al., 2023). On the other hand, the processing pipelines we have employed are fully open-source, and we encourage further exploration of multimodal approaches using datasets that are more homogeneous or specifically designed to minimize inter-site variability.

## Conclusion

Our findings demonstrate that neuroimaging and neuropsychological features play distinct roles in age prediction and disease classification in the context of BrainAge modeling, with neuroimaging features excelling in age prediction and neuropsychological features showing greater sensitivity to clinical distinctions. The hippocampus and grey matter volume emerge as critical biomarkers in both aging and Alzheimer’s disease progression. The instability of results across different model configurations and data splits suggests a need for further exploration into feature robustness and optimal model selection based on task-specific requirements. Overall, while BrainAge deltas can provide a useful single metric associated with the risk of developing Alzheimer’s disease, their application should be carefully considered in the context of specific clinical and research goals.

## Data Availability

Data used in preparation of this article were obtained from the Alzheimers Disease Neuroimaging Initiative (ADNI) database (adni.loni.usc.edu). As such, the investigators within the ADNI contributed to the design and implementation of ADNI and/or provided data but did not participate in the analysis or the writing of this report. A complete listing of ADNI investigators can be found in link

https://adni.loni.usc.edu/wp-content/uploads/how_to_apply/ADNI_Acknowledgement_List.pdf

## Ethic statement

Not applicable. Data was obtained from the Alzheimer’s Neuroimaging Data Initiative (ADNI) see www.adni.loni.usc.edu.

## CRediT authorship

**Jorge Garcia Condado**: Conceptualization, Methodology, Data curation, Formal analysis, Visualization, Writing – original draft, Writing – review & editing. **Ines Verdugo Recuerdo:** Methodology, Formal analysis, Visualization, Writing – review & editing. **Iñigo Tellaetxe Elorriaga**: Data curation, Visualization, Writing – original draft **Colin Birkhenbil:** Writing – review & editing. **Maria Carrigan**: Writing – review & editing **Ibai Diez**: Writing – review & editing. **Rachel F. Buckley**: Writing – review & editing. **Asier Erramuzpe:** Supervision, Project administration, Writing – review & editing. **Jesus M. Cortes**: Supervision, Project administration, Writing – review & editing.

## Acknowledgments

Data used in preparation of this article were obtained from the Alzheimer’s Disease Neuroimaging Initiative (ADNI) database (adni.loni.usc.edu). As such, the investigators within the ADNI contributed to the design and implementation of ADNI and/or provided data but did not participate in the analysis or the writing of this report. A complete listing of ADNI investigators can be found here.

We want to thank Vanessa Goméz Verdejo from Universidad Carlos III for her valuable comments on the work.

The project that gave rise to these results received the support of a fellowship from “la Caixa” Foundation (ID 100010434) to J.G.C. The fellowship code is LCF/BQ/DI21/11860030. J.M.C acknowledges financial support from the Health Department of the Basque Country (grants 202211103 and 2023111002). A.E. is supported by the Spanish Ministry of Science and Innovation, grant RYC2021-032390-I. J.M.C. and A.E. are supported by Ikerbasque: The Basque Foundation for Science and the Spanish Ministry of Science under Grant PID2023-148012OB-I00.

Data collection and sharing for this project was funded by the Alzheimer’s Disease Neuroimaging Initiative (ADNI) (National Institutes of Health Grant U01 AG024904) and DOD ADNI (Department of Defense award number W81XWH-12-2-0012). ADNI is funded by the National Institute on Aging, the National Institute of Biomedical Imaging and Bioengineering, and through generous contributions from the following: AbbVie, Alzheimer’s Association; Alzheimer’s Drug Discovery Foundation; Araclon Biotech; BioClinica, Inc.; Biogen; Bristol-Myers Squibb Company; CereSpir, Inc.; Cogstate; Eisai Inc.; Elan Pharmaceuticals, Inc.; Eli Lilly and Company; EuroImmun; F. Hoffmann-La Roche Ltd and its affiliated company Genentech, Inc.; Fujirebio; GE Healthcare; IXICO Ltd.; Janssen Alzheimer Immunotherapy Research & Development, LLC.; Johnson & Johnson Pharmaceutical Research & Development LLC.; Lumosity; Lundbeck; Merck & Co., Inc.; Meso Scale Diagnostics, LLC.; NeuroRx Research; Neurotrack Technologies; Novartis Pharmaceuticals Corporation; Pfizer Inc.; Piramal Imaging; Servier; Takeda Pharmaceutical Company; and Transition Therapeutics. The Canadian Institutes of Health Research is providing funds to support ADNI clinical sites in Canada. Private sector contributions are facilitated by the Foundation for the National Institutes of Health (www.fnih.org). The grantee organization is the Northern California Institute for Research and Education, and the study is coordinated by the Alzheimer’s Therapeutic Research Institute at the University of Southern California. ADNI data are disseminated by the Laboratory for Neuro Imaging at the University of Southern California.

## Code Availability

The software used in this work is open source and publicly available: www.github.com/compneurobilbao/ageml.

## Supplementary Figures

**Supplementary Figure 1.**
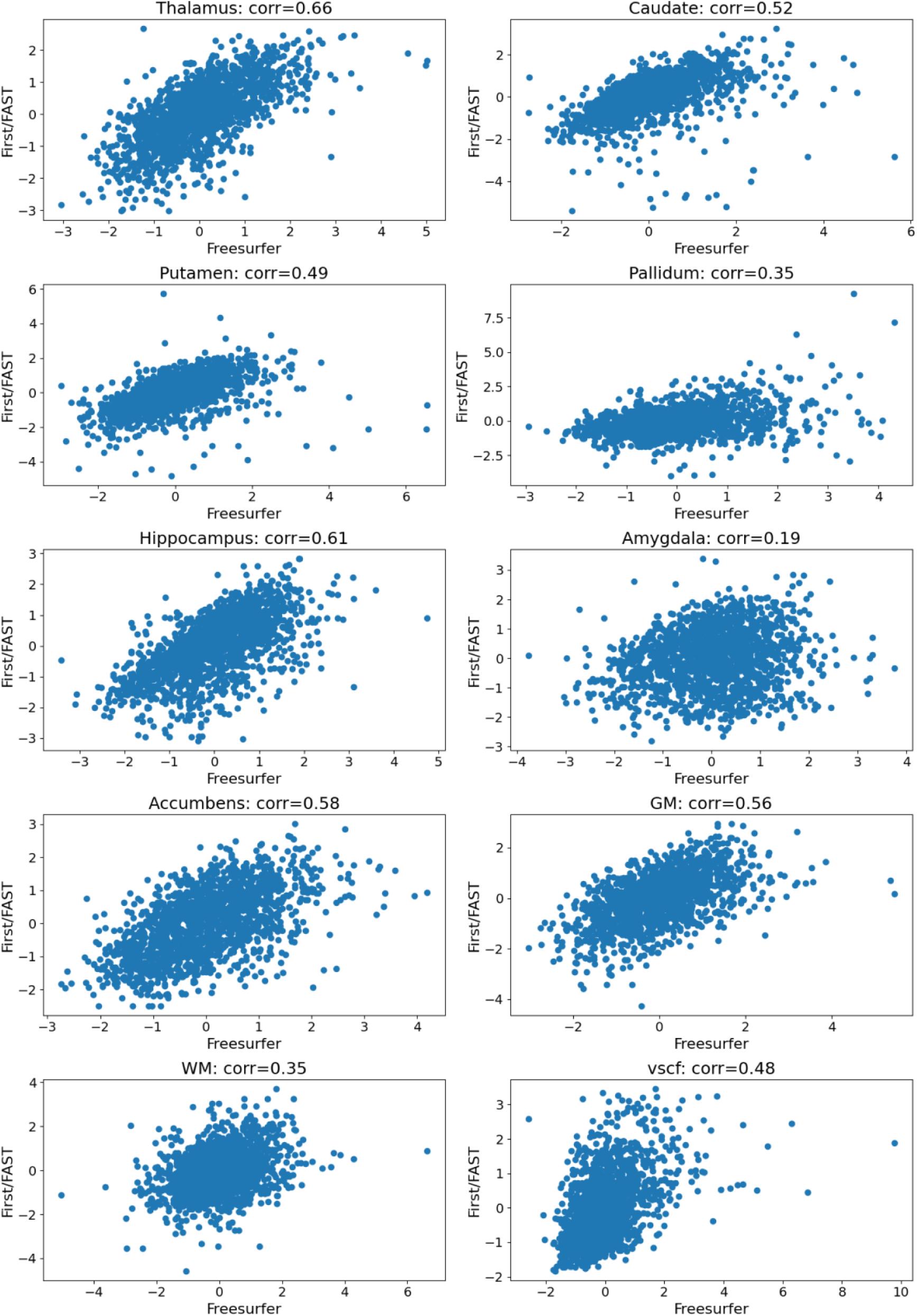
Normalized values for FIRST/FAST segmentation of cortical and subcortical volumes in comparison to normalized values of FreeSurfer Segmentation. Values shown are Pearson correlation values.

**Supplementary Figure 2.**
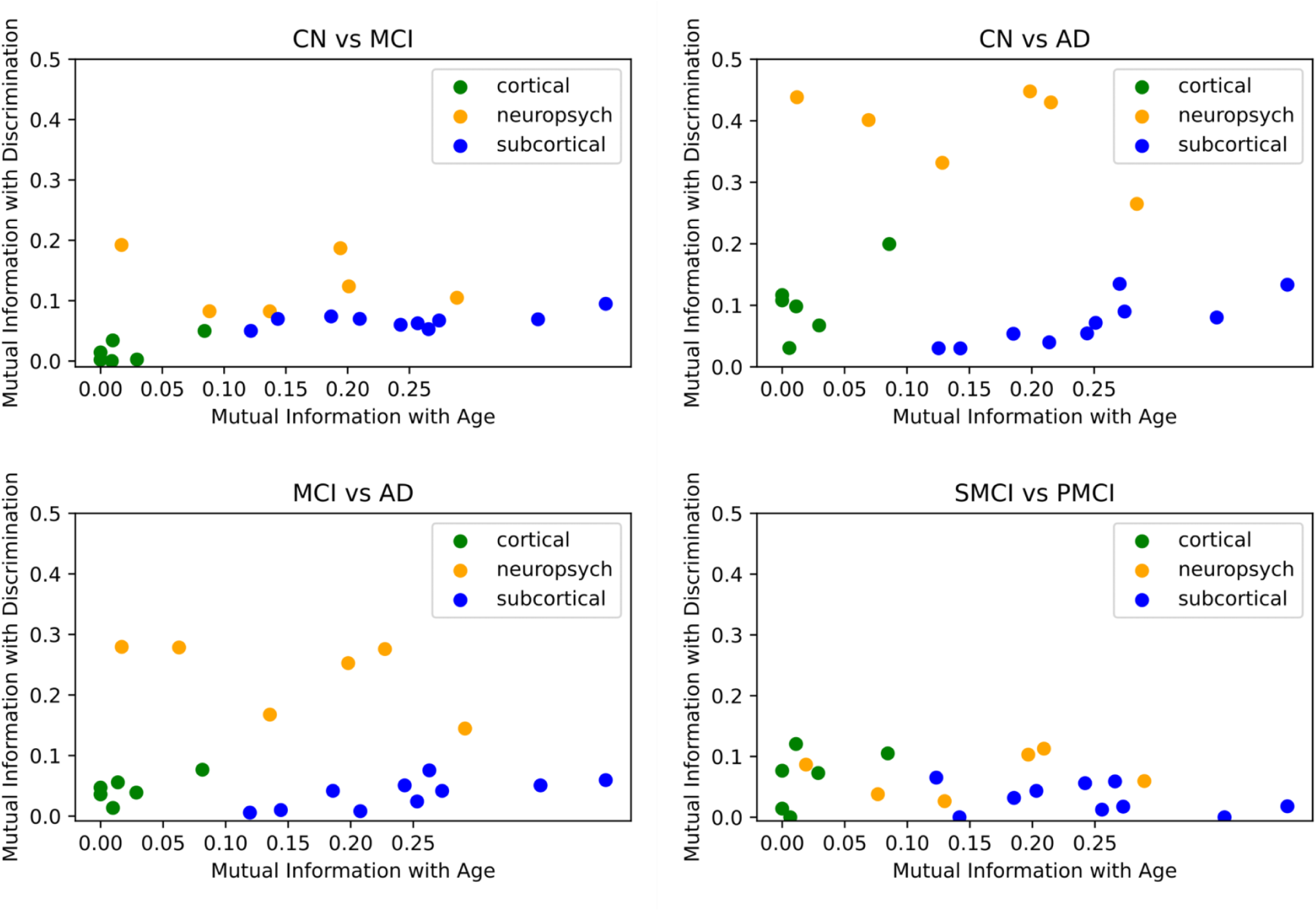
Mutual information of each feature with Age and their discriminative power. In blue are features derived from neuroimaging metrics of subcortical volumes using FIRST/FAST, in orange features derived from neuropsychological tests and in green neuroimaging metrics of cortical thickness using FreeSurfer. Control (CN), Alzheimer’s Disease (AD), Mild Cognitive Impairment (MCI), stable MCI (sMCI), and progressive MCI (pMCI). Each specific feature name in the graph can be derived from Supplementary Table 1.

**Supplementary Figure. 3.**
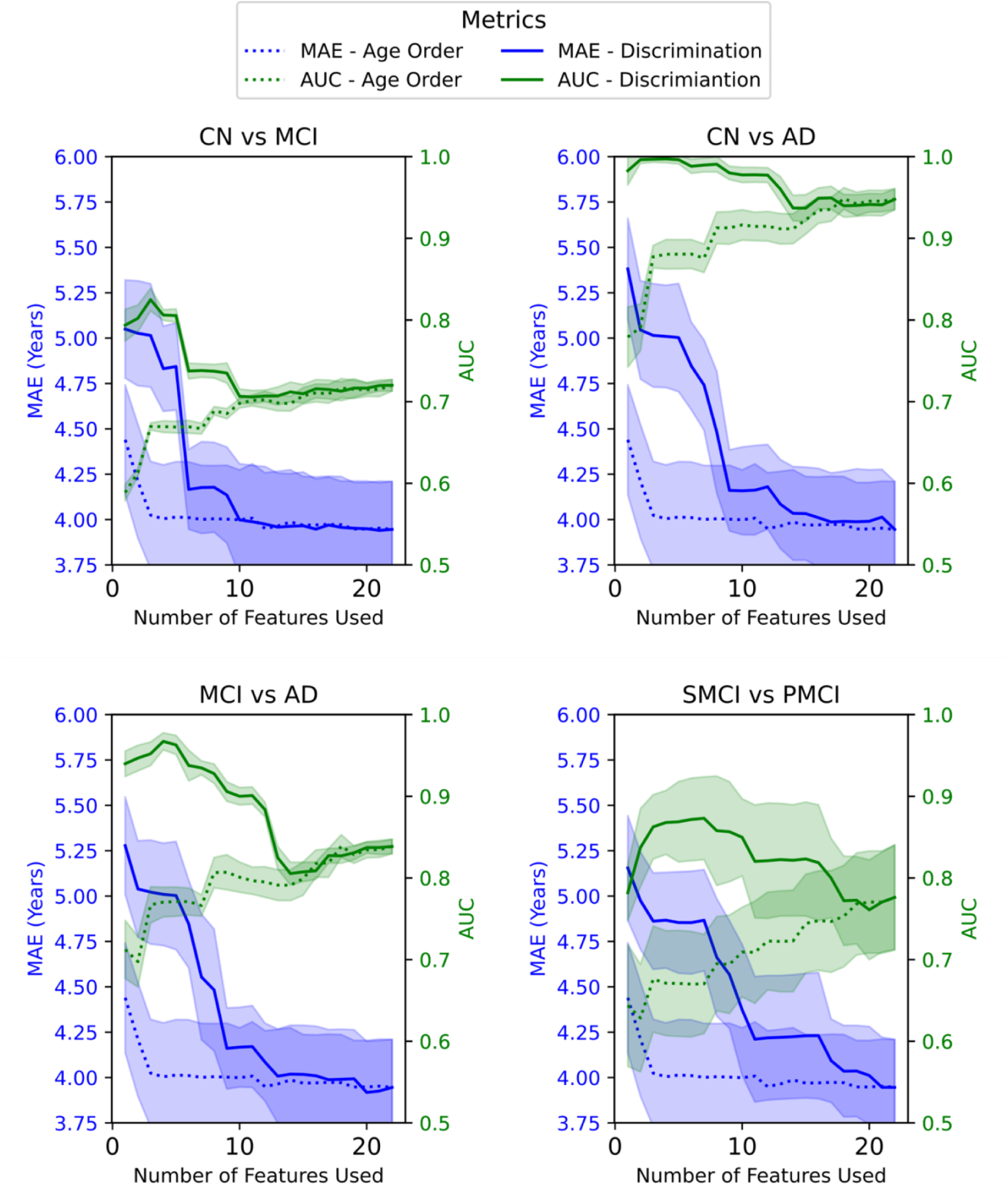
Comparison of the performance metrics, Mean Absolute Error (MAE), and Area Under the Curve (AUC) for different health condition groups using BrainAge Modelling including cortical thickness measurements. Features are added to the BrainAge model in descending order based on their age relationship (dotted line). Additionally, features are added to the BrainAge model in descending order according to their importance in discriminating between the following groups: Control (CN), Alzheimer’s Disease (AD), Mild Cognitive Impairment (MCI), stable MCI (sMCI), and progressive MCI (pMCI) (solid line). The shaded areas show the 95% confidence intervals of the MAE and AUC measurements.

**Figure. 4.**
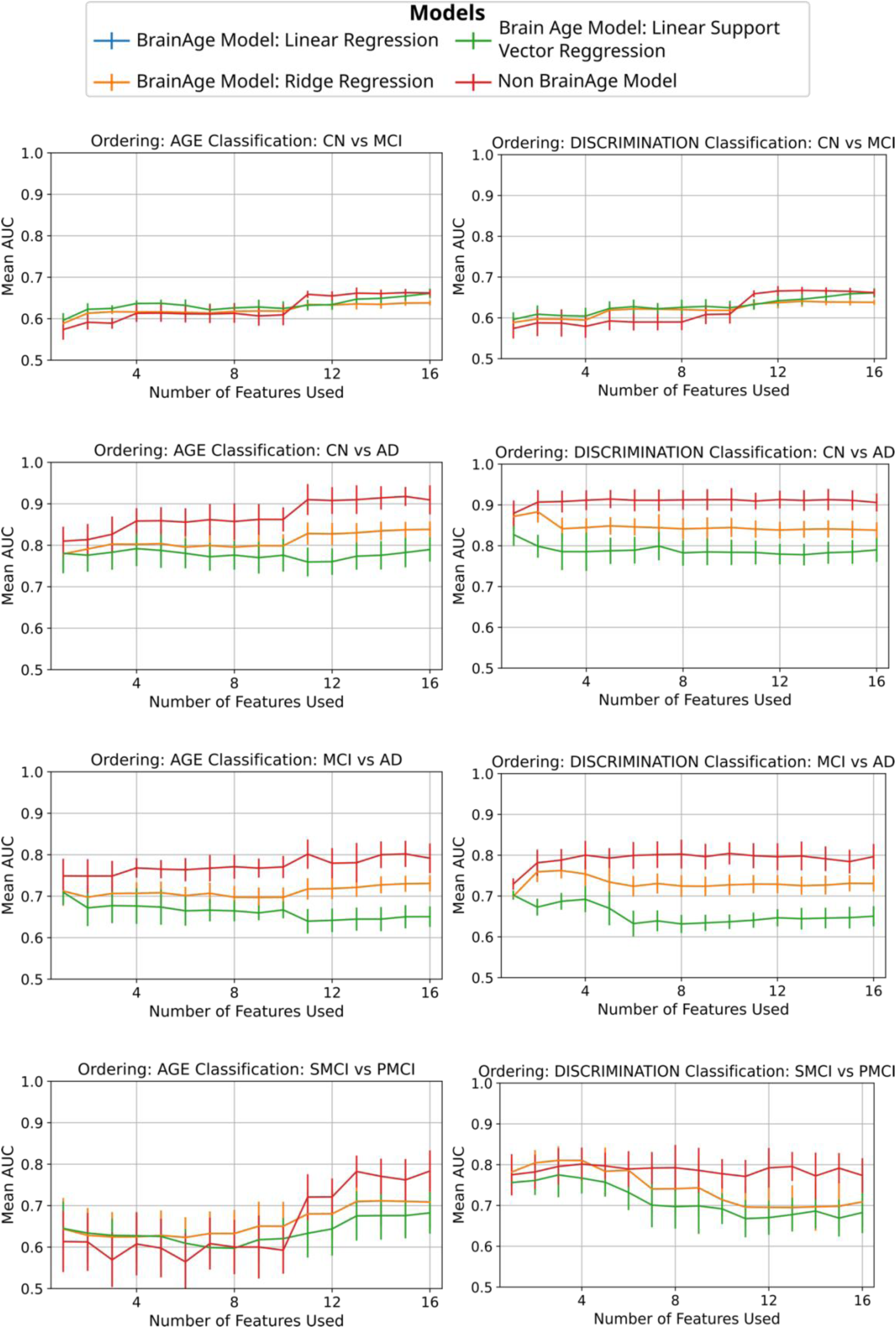
Comparison of the Area Under the Curve (AUC) using different machine learning models for classification using neuroimaging features including cortical thickness measurements. *Blue line:* Input to logistic regressor: Delta. BrainAge model: linear regression. *Orange line:* Input to logistic regressor: Delta. BrainAge model: Ridge. *Green line:* Input to logistic regressor: Delta. BrainAge model: Support Vector Regressor. *Red line:* Input to logistic regressor: Neuroimaging Features, No BrainAge modeling. Models are tested and trained across clinical classification groups and features ordering by Age Relationship and by discrimination between groups order (Control Group (CN), Alzheimer Disease (AD), Mild Cognitive Impairment (MCI), stable Mild Cognitive Impairment (sMCI) and progressive Mild Cognitive Impairment (pMCI)). The blue and orange line overlap Error bars show the standard deviation of the AUC across CV folds.

## Supplementary Tables

**Table 1.** Results of ranking features across different orderings including neuroimaging volumetric features, neuroimaging cortical thickness features and neuropsychological features. The features are ordered in descending importance according to the variable indicated in the column header. The first column indicates ordering according to mutual information with age, while the subsequent columns make a comparison between the mutual information of features for the following groups: Control Group (CN), Alzheimer’s Diseases (AD), Mild Cognitive Impairment (MCI), Stable Mild Cognitive Impairment (sMCI) and Progressive Mild Cognitive Impairment (pMCI).

| Set | Age | CN vs AD | CN vs MCI | MCI vs AD | sMCI vs pMCI |
| --- | --- | --- | --- | --- | --- |
| 1 | Grey Matter | FAQ | ADNI Memory | MMSE | Fusiform |
| 2 | Thalamus | ADNI Memory | ADAS | ADNI Memory | ADAS |
| 3 | ADNI Executive Function | ADAS | FAQ | ADAS | ADNI Memory |
| 4 | Cerebrospinal Fluid | MMSE | ADNI Executive Function | FAQ | FAQ |
| 5 | Hippocampus | MoCA | MoCA | MoCA | Entorhinal |
| 6 | Putamen | ADNI Executive Function | Grey Matter | ADNI Executive Function | Parahippocampal |
| 7 | Accumbens | Entorhinal | Amygdala | Hippocampus | Inferior temporal |
| 8 | ADAS | Hippocampus | Pallidum | Entorhinal | White Matter |
| 9 | Pallidum | Grey Matter | Caudate | Grey Matter | ADNI Executive Function |
| 10 | ADNI Memory | Inferior temporal | Thalamus | Fusiform | Hippocampus |
| 11 | Amygdala | Middle temporal | Cerebrospinal Fluid | Middle temporal | Accumbens |
| 12 | Caudate | Fusiform | Putamen | Accumbens | Pallidum |
| 13 | MoCA | Cerebrospinal Fluid | Accumbens | Thalamus | MMSE |
| 14 | White Matter | Thalamus | MMSE | Amygdala | MoCA |
| 15 | Entorhinal | Putamen | Hippocampus | Cerebrospinal Fluid | Amygdala |
| 16 | MMSE | Parahippocampal | Entorhinal | Inferior temporal | Middle temporal |
| 17 | Parahippocampal | Accumbens | White Matter | Parahippocampal | Grey Matter |
| 18 | FAQ | Amygdala | Fusiform | Putamen | Cerebrospinal Fluid |
| 19 | Fusiform | Pallidum | Inferior temporal | Inferior parietal | Putamen |
| 20 | Inferior parietal | Inferior parietal | Inferior parietal | Caudate | Thalamus |
| 21 | Middle temporal | White Matter | Parahippocampal | Pallidum | Caudate |
| 22 | Inferior temporal | Caudate | Middle temporal | White Matter | Inferiorparietal |

**Supplementary Table 2.**
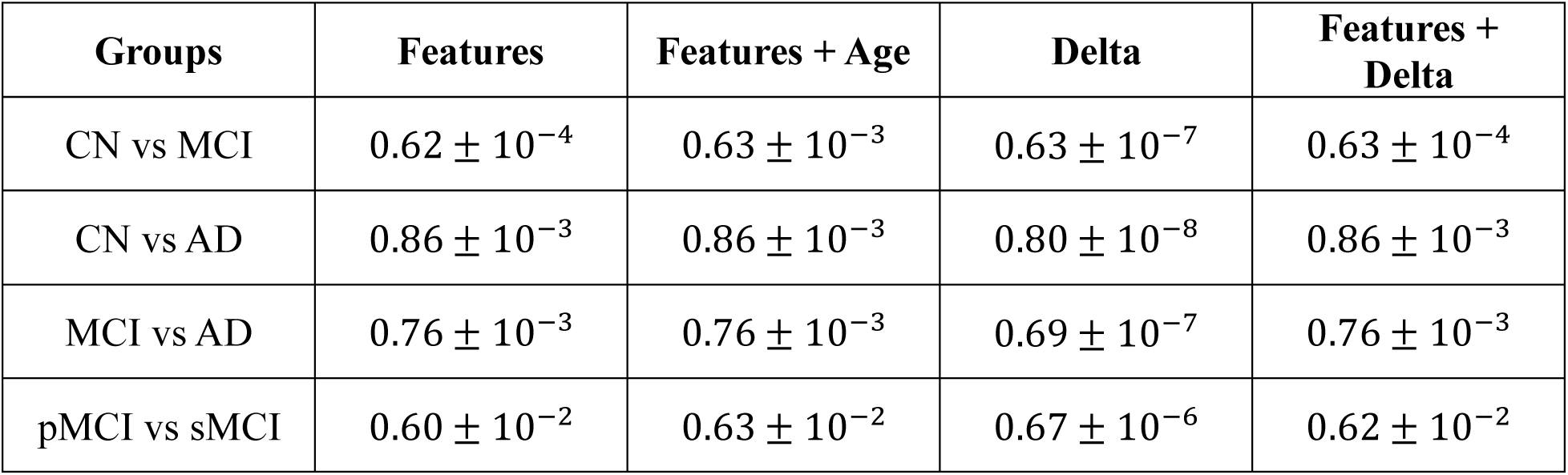
Comparison of the Area Under the Curve (AUC) across various clinical classifications using different input feature sets and 5 different random seeds. . The classification was performed using a Logistic Regressor and included all brain structural features for the following groups: Control Group (CN), Alzheimer’s Disease (AD), Mild Cognitive Impairment (MCI), stable Mild Cognitive Impairment (sMCI), and progressive Mild Cognitive Impairment (pMCI). ± indicates standard deviations across seeds.

| **Groups** | **Features** | **Features + Age** | **Delta** | **Features + Delta** |
| --- | --- | --- | --- | --- |
| CN vs MCI | $0.62 \pm 10^{-4}$ | $0.63 \pm 10^{-3}$ | $0.63 \pm 10^{-7}$ | $0.63 \pm 10^{-4}$ |
| CN vs AD | $0.86 \pm 10^{-3}$ | $0.86 \pm 10^{-3}$ | $0.80 \pm 10^{-8}$ | $0.86 \pm 10^{-3}$ |
| MCI vs AD | $0.76 \pm 10^{-3}$ | $0.76 \pm 10^{-3}$ | $0.69 \pm 10^{-7}$ | $0.76 \pm 10^{-3}$ |
| pMCI vs sMCI | $0.60 \pm 10^{-2}$ | $0.63 \pm 10^{-2}$ | $0.67 \pm 10^{-6}$ | $0.62 \pm 10^{-2}$ |

## References

A new rating scale for Alzheimer’s disease, 1984. AJP 141, 1356–1364. 10.1176/ajp.141.11.1356

Baecker, L., Garcia-Dias, R., Vieira, S., Scarpazza, C., Mechelli, A., 2021. Machine learning for brain age prediction: Introduction to methods and clinical applications. eBioMedicine 72, 103600. 10.1016/j.ebiom.2021.103600

Bakkour, A., Morris, J.C., Dickerson, B.C., 2009. The cortical signature of prodromal AD: Regional thinning predicts mild AD dementia. Neurology 72, 1048–1055. 10.1212/01.wnl.0000340981.97664.2f

Bonifazi, P., Erramuzpe, A., Diez, I., Gabilondo, I., Boisgontier, M.P., Pauwels, L., Stramaglia, S., Swinnen, S.P., Cortes, J.M., 2018. Structure–function multi-scale connectomics reveals a major role of the fronto-striato-thalamic circuit in brain aging. Human Brain Mapping 39, 4663–4677. 10.1002/hbm.24312

de Lange, A.G., Anatürk, M., Rokicki, J., Han, L.K.M., Franke, K., Alnæs, D., Ebmeier, K.P., Draganski, B., Kaufmann, T., Westlye, L.T., Hahn, T., Cole, J.H., 2022. Mind the gap: Performance metric evaluation in brain-age prediction. Human Brain Mapping 43, 3113– 3129. 10.1002/hbm.25837

de Lange, A.-M.G., Cole, J.H., 2020. Commentary: Correction procedures in brain-age prediction. NeuroImage: Clinical 26, 102229. 10.1016/j.nicl.2020.102229

Dular, L., Špiclin, Z., 2024. BASE: Brain Age Standardized Evaluation. NeuroImage 285, 120469. 10.1016/j.neuroimage.2023.120469

Fischl, B., 2012. FreeSurfer. NeuroImage 62, 774–781. 10.1016/j.neuroimage.2012.01.021

Folstein, M.F., Folstein, S.E., McHugh, P.R., 1975. “Mini-mental state.” Journal of Psychiatric Research 12, 189–198. 10.1016/0022-3956(75)90026-6

for the Alzheimer’s Disease Neuroimaging Initiative, Crane, P.K., Carle, A., Gibbons, L.E., Insel, P., Mackin, R.S., Gross, A., Jones, R.N., Mukherjee, S., Curtis, S.M., Harvey, D., Weiner, M., Mungas, D., 2012a. Development and assessment of a composite score for memory in the Alzheimer’s Disease Neuroimaging Initiative (ADNI). Brain Imaging and Behavior 6, 502–516. 10.1007/s11682-012-9186-z

for the Alzheimer’s Disease Neuroimaging Initiative, Gibbons, L.E., Carle, A.C., Mackin, R.S., Harvey, D., Mukherjee, S., Insel, P., Curtis, S.M., Mungas, D., Crane, P.K., 2012b. A composite score for executive functioning, validated in Alzheimer’s Disease Neuroimaging Initiative (ADNI) participants with baseline mild cognitive impairment. Brain Imaging and Behavior 6, 517–527. 10.1007/s11682-012-9176-1

Franke, K., Gaser, C., 2019. Ten Years of BrainAGE as a Neuroimaging Biomarker of Brain Aging: What Insights Have We Gained? Front. Neurol. 10, 789. 10.3389/fneur.2019.00789

Franke, K., Ziegler, G., Klöppel, S., Gaser, C., 2010. Estimating the age of healthy subjects from T1-weighted MRI scans using kernel methods: Exploring the influence of various parameters. NeuroImage 50, 883–892. 10.1016/j.neuroimage.2010.01.005

Garcia Condado, J., Cortes, J.M., for the Alzheimer’s Disease Neuroimaging Initiative, 2023. NeuropsychBrainAge: A biomarker for conversion from mild cognitive impairment to Alzheimer’s disease. Alz & Dem Diag Ass & Dis Mo 15, e12493. 10.1002/dad2.12493

Garcia Condado, J., Elorriaga, I.T., Cortes, J.M., Erramuzpe, A., 2025. AgeML: Age modeling with Machine Learning. IEEE J. Biomed. Health Inform. 1–11. 10.1109/JBHI.2025.3531017

Gaser, C., Franke, K., Klöppel, S., Koutsouleris, N., Sauer, H., Alzheimer’s Disease Neuroimaging Initiative, 2013. BrainAGE in Mild Cognitive Impaired Patients: Predicting the Conversion to Alzheimer’s Disease. PLoS ONE 8, e67346. 10.1371/journal.pone.0067346

Gebre, R.K., Senjem, M.L., Raghavan, S., Schwarz, C.G., Gunter, J.L., Hofrenning, E.I., Reid, R.I., Kantarci, K., Graff-Radford, J., Knopman, D.S., Petersen, R.C., Jack, C.R., Vemuri, P., 2023. Cross–scanner harmonization methods for structural MRI may need further work: A comparison study. NeuroImage 269, 119912. 10.1016/j.neuroimage.2023.119912

Guan, S., Jiang, R., Meng, C., Biswal, B., 2023. Brain age prediction across the human lifespan using multimodal MRI data. GeroScience 46, 1–20. 10.1007/s11357-023-00924-0

Hwang, G., Abdulkadir, A., Erus, G., Habes, M., Pomponio, R., Shou, H., Doshi, J., Mamourian, E., Rashid, T., Bilgel, M., Fan, Y., Sotiras, A., Srinivasan, D., Morris, J.C., Albert, M.S., Bryan, N.R., Resnick, S.M., Nasrallah, I.M., Davatzikos, C., Wolk, D.A., from the iSTAGING consortium, for the ADNI, 2022. Disentangling Alzheimer’s disease neurodegeneration from typical brain ageing using machine learning. Brain Communications 4, fcac117. 10.1093/braincomms/fcac117

Korbmacher, M., Wang, M.-Y., Eikeland, R., Buchert, R., Leonardsen, E., Westlye, L.T., Maximov, I.I., Specht, K., 2023. Brain age predictions in longitudinal data reveal the importance of scan quality and field strength (preprint). Neuroscience. 10.1101/2023.03.31.535038

Moguilner, S., Baez, S., Hernandez, H., Migeot, J., Legaz, A., Gonzalez-Gomez, R., Farina, F.R., Prado, P., Cuadros, J., Tagliazucchi, E., Altschuler, F., Maito, M.A., Godoy, M.E., Cruzat, J., Valdes-Sosa, P.A., Lopera, F., Ochoa-Gómez, J.F., Hernandez, A.G., Bonilla-Santos, J., Gonzalez-Montealegre, R.A., Anghinah, R., d’Almeida Manfrinati, L.E., Fittipaldi, S., Medel, V., Olivares, D., Yener, G.G., Escudero, J., Babiloni, C., Whelan, R., Güntekin, B., Yırıkoğulları, H., Santamaria-Garcia, H., Lucas, A.F., Huepe, D., Di Caterina, G., Soto-Añari, M., Birba, A., Sainz-Ballesteros, A., Coronel-Oliveros, C., Yigezu, A., Herrera, E., Abasolo, D., Kilborn, K., Rubido, N., Clark, R.A., Herzog, R., Yerlikaya, D., Hu, K., Parra, M.A., Reyes, P., García, A.M., Matallana, D.L., Avila-Funes, J.A., Slachevsky, A., Behrens, M.I., Custodio, N., Cardona, J.F., Barttfeld, P., Brusco, I.L., Bruno, M.A., Sosa Ortiz, A.L., Pina-Escudero, S.D., Takada, L.T., Resende, E., Possin, K.L., De Oliveira, M.O., Lopez-Valdes, A., Lawlor, B., Robertson, I.H., Kosik, K.S., Duran-Aniotz, C., Valcour, V., Yokoyama, J.S., Miller, B., Ibanez, A., 2024. Brain clocks capture diversity and disparities in aging and dementia across geographically diverse populations. Nat Med. 10.1038/s41591-024-03209-x

Mu, Y., Gage, F.H., 2011. Adult hippocampal neurogenesis and its role in Alzheimer’s disease. Mol Neurodegeneration 6, 85. 10.1186/1750-1326-6-85

Nasreddine, Z.S., Phillips, N.A., Bédirian, V., Charbonneau, S., Whitehead, V., Collin, I., Cummings, J.L., Chertkow, H., 2005. The Montreal Cognitive Assessment, MoCA: A Brief Screening Tool For Mild Cognitive Impairment: MOCA: A BRIEF SCREENING TOOL FOR MCI. Journal of the American Geriatrics Society 53, 695–699. 10.1111/j.1532-5415.2005.53221.x

Oh, H.S.-H., Rutledge, J., Nachun, D., Pálovics, R., Abiose, O., Moran-Losada, P., Channappa, D., Urey, D.Y., Kim, K., Sung, Y.J., Wang, L., Timsina, J., Western, D., Liu, M., Kohlfeld, P., Budde, J., Wilson, E.N., Guen, Y., Maurer, T.M., Haney, M., Yang, A.C., He, Z., Greicius, M.D., Andreasson, K.I., Sathyan, S., Weiss, E.F., Milman, S., Barzilai, N., Cruchaga, C., Wagner, A.D., Mormino, E., Lehallier, B., Henderson, V.W., Longo, F.M., Montgomery, S.B., Wyss-Coray, T., 2023. Organ aging signatures in the plasma proteome track health and disease. Nature 624, 164–172. 10.1038/s41586-023-06802-1

Patenaude, B., Smith, S.M., Kennedy, D.N., Jenkinson, M., 2011. A Bayesian model of shape and appearance for subcortical brain segmentation. NeuroImage 56, 907–922. 10.1016/j.neuroimage.2011.02.046

Petersen, R.C., Aisen, P.S., Beckett, L.A., Donohue, M.C., Gamst, A.C., Harvey, D.J., Jack, C.R., Jagust, W.J., Shaw, L.M., Toga, A.W., Trojanowski, J.Q., Weiner, M.W., 2010. Alzheimer’s Disease Neuroimaging Initiative (ADNI): Clinical characterization. Neurology 74, 201–209. 10.1212/WNL.0b013e3181cb3e25

Pfeffer, R.I., Kurosaki, T.T., Harrah, C.H., Chance, J.M., Filos, S., 1982. Measurement of Functional Activities in Older Adults in the Community. Journal of Gerontology 37, 323–329. 10.1093/geronj/37.3.323

Pichet Binette, A., Gonneaud, J., Vogel, J.W., La Joie, R., Rosa-Neto, P., Collins, D.L., Poirier, J., Breitner, J.C.S., Villeneuve, S., Vachon-Presseau, E., for the Alzheimer’s Disease Neuroimaging Initiative, the PREVENT-AD Research Group, 2020. Morphometric network differences in ageing versus Alzheimer’s disease dementia. Brain 143, 635–649. 10.1093/brain/awz414

Poulin, S.P., Dautoff, R., Morris, J.C., Barrett, L.F., Dickerson, B.C., 2011. Amygdala atrophy is prominent in early Alzheimer’s disease and relates to symptom severity. Psychiatry Research: Neuroimaging 194, 7–13. 10.1016/j.pscychresns.2011.06.014

Puc, A., Struc, V., Grm, K., 2021. Analysis of Race and Gender Bias in Deep Age Estimation Models, in: 2020 28th European Signal Processing Conference (EUSIPCO). Presented at the 2020 28th European Signal Processing Conference (EUSIPCO), IEEE, Amsterdam, Netherlands, pp. 830–834. 10.23919/Eusipco47968.2020.9287219

Sabuncu, M.R., 2011. The Dynamics of Cortical and Hippocampal Atrophy in Alzheimer Disease. Arch Neurol 68, 1040. 10.1001/archneurol.2011.167

Scikit-learn: Machine Learning in Python, n.d. 2011 12, 2825–2830.

Smith, S.M., Jenkinson, M., Woolrich, M.W., Beckmann, C.F., Behrens, T.E.J., Johansen-Berg, H., Bannister, P.R., De Luca, M., Drobnjak, I., Flitney, D.E., Niazy, R.K., Saunders, J., Vickers, J., Zhang, Y., De Stefano, N., Brady, J.M., Matthews, P.M., 2004. Advances in functional and structural MR image analysis and implementation as FSL. NeuroImage 23, S208–S219. 10.1016/j.neuroimage.2004.07.051

Smith, S.M., Zhang, Y., Jenkinson, M., Chen, J., Matthews, P.M., Federico, A., De Stefano, N., 2002. Accurate, Robust, and Automated Longitudinal and Cross-Sectional Brain Change Analysis. NeuroImage 17, 479–489. 10.1006/nimg.2002.1040

Tan, T.W.K., Nguyen, K., Zhang, C., Kong, R., Cheng, S.F., Ji, F., Chong, J.S.X., Chong, E.J.Y., Venketasubramanian, N., Orban, C., Chee, M.W.L., Chen, C., Zhou, J.H., Thomas Yeo, B.T., The Alzheimer’s Disease Neuroimaging Initiative, The Australian Imaging Biomarkers and Lifestyle Study of Aging, 2025. Mind the Gap: Does Brain Age Improve Alzheimer’s Disease Prediction? Human Brain Mapping 46, e70276. 10.1002/hbm.70276

Warrington, S., Torchi, A., Mougin, O., Campbell, J., Ntata, A., Craig, M., Assimopoulos, S., Alfaro-Almagro, F., Miller, K.L., Jenkinson, M., Morgan, P.S., Sotiropoulos, S.N., 2025. A multi-site, multi-modal travelling-heads resource for brain MRI harmonisation. Sci Data 12, 609. 10.1038/s41597-025-04822-2

Yu, Y., Cui, H., Haas, S.S., New, F., Sanford, N., Yu, K., Zhan, D., Yang, G., Gao, J., Wei, D., Qiu, J., Banaj, N., Boomsma, D.I., Breier, A., Brodaty, H., Buckner, R.L., Buitelaar, J.K., Cannon, D.M., Caseras, X., Clark, V.P., Conrod, P.J., Crivello, F., Crone, E.A., Dannlowski, U., Davey, C.G., de Haan, L., de Zubicaray, G.I., Di Giorgio, A., Fisch, L., Fisher, S.E., Franke, B., Glahn, D.C., Grotegerd, D., Gruber, O., Gur, R.E., Gur, R.C., Hahn, T., Harrison, B.J., Hatton, S., Hickie, I.B., Hulshoff Pol, H.E., Jamieson, A.J., Jernigan, T.L., Jiang, J., Kalnin, A.J., Kang, S., Kochan, N.A., Kraus, A., Lagopoulos, J., Lazaro, L., McDonald, B.C., McDonald, C., McMahon, K.L., Mwangi, B., Piras, F., Rodriguez-Cruces, R., Royer, J., Sachdev, P.S., Satterthwaite, T.D., Saykin, A.J., Schumann, G., Sevaggi, P., Smoller, J.W., Soares, J.C., Spalletta, G., Tamnes, C.K., Trollor, J.N., Van’t Ent, D., Vecchio, D., Walter, H., Wang, Y., Weber, B., Wen, W., Wierenga, L.M., Williams, S.C.R., Wu, M., Zunta-Soares, G.B., Bernhardt, B., Thompson, P., Frangou, S., Ge, R., 2024. Brain-age prediction: Systematic evaluation of site effects, and sample age range and size. Hum Brain Mapp 45, e26768. 10.1002/hbm.26768

